# Von Meyenburg Complexes and Malignancy: A Systematic Review

**DOI:** 10.1101/2021.04.17.21255674

**Authors:** Abu Baker Sheikh, Anthony Nguyen, Abdul Ahad Ehsan Sheikh, Nismat Javed, Katarina Leyba, Rahul Shekhar

## Abstract

Von Meyenburg Complexes (VMC) are benign lesions secondary to ductal plate malformations. These are relatively rare in adults and are often asymptomatic, with many lesions being diagnosed on imaging. Despite these benign characteristics, VMC have the potential to undergo malignant changes leading to conditions such as cholangiocarcinoma and other cancers. The objective of the review is to report and analyze the cases of biliary hamartomas which were associated with malignancy. We performed a structured systematic review of literature and identified 31 cases of biliary duct hamartomas associated with malignancy. The mean age at onset was 61 years±13 (min 19 to max 88) with the majority of the patients being male (64.5%). Of all cases, 41.9% reported symptoms (13/31 cases) with the most common symptom being abdominal pain (32.3%). Biopsies reported malignancy in 77.4% of cases (24/31 cases) with the most common reported associated malignancy being cholangiocarcinoma 54.8% (17/31). 93.3% of the cases underwent surgical intervention (28/30 cases). VMC are an important entity with malignancy potential which is under-reported. Presence of ‘red flag’ symptoms such as weight loss and obstructive jaundice in patients with diagnosed biliary hamartomas should prompt an evaluation for malignancy.

## INTRODUCTION

Biliary Hamartomas, also referred to as Von Meyenburg Complexes (VMC), are hepatic tumor like lesions that form as a result of ductal plate malformations. VMCs have an incidence of approximately 5.6% in adults [1], are often subclinical and are most often found incidentally on abdominal CT or MRI. Histologically, they are characterized by dilated and irregularly formed bile ducts with bland cuboidal epithelium. Although most often asymptomatic, the clinical relevance of VMCs lie in their potential to transform into more malignant pathologies such as intrahepatic cholangiocarcinoma (ICC) and less often, hepatocellular carcinoma (HCC). There have been numerous case reports of VMCs with subsequent transformation into ICC or HCC however the actual rate of transformation into malignancy and its type is unclear at this time. This is a systematic review that compiles 31 cases of biliary hamartomas with associated malignancy dating back to 1975.

## METHODS

### Protocol development and systematic review registration

We developed the protocol for this review after consensus with all the reviewers and subject experts. After protocol preparation, data search was done. We kept our data search broad.

### Search strategy

Keywords (including all commonly used abbreviations of these terms) used in the search strategy were as follows: “Biliary hamartomas” OR “Von Meyenburg Complexes” OR “Biliary duct hamartomas” OR “Hamartomas AND biliary” OR “Multiple biliary hamartomas” OR “Multiple biliary duct hamartomas”.

### Data extraction (selection and coding)

We screened PubMed, Web of Science and CINAHL databases and articles published in the English language were included in the systemic review. On the initial search, 163 articles on PubMed, 33 on Web of Science (excluding Medline) and 63 on CINAHL (excluding Medline) were included. After the initial search, duplicates were removed and we imported all included search study into EndNote online software. Two independent reviewers screened remaining studies for the inclusion based on inclusion criteria, and researchers were blinded to each other’s decisions. Rayyan software and Mendeley desktop were used. Once the initial screening is done, two independent reviewers reviewed the full-text article for final inclusion. Reviewers were blinded to each other’s decisions, and a third reviewer resolved any dispute.

Data was extracted from study documents, including information about study design and methodology, participant demographics and baseline characteristics, publication journal, clinical presentation, symptoms, laboratory data, imaging data, intervention, treatment, clinical outcomes, morbidity and mortality.

One reviewer did data extraction, and another reviewer cross-checked the extracted data for accuracy and completeness. Publications that were not peer-reviewed were excluded from this study. The preliminary data was entered and recorded in an excel spreadsheet. The final analysis included 31 studies (figure 1).

**Figure 1:**
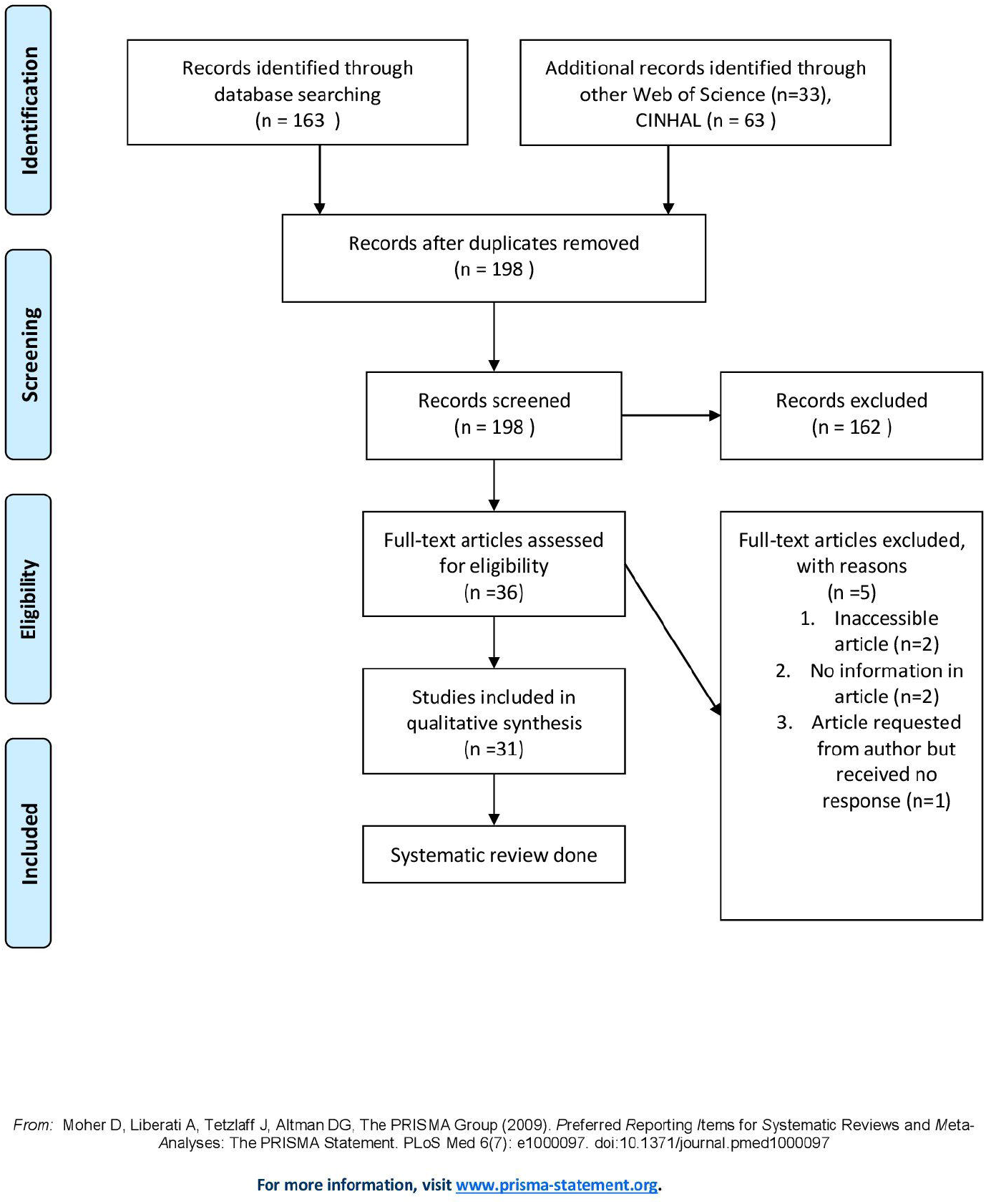
PRISMA 2009 Flow Diagram.

### Strategy for data synthesis

For statistical analysis, we used IBM SPSS Statistics version 21 (IBM, Armonk, NY, USA). Based on the distribution of values, continuous data were expressed as mean ± standard deviation. Qualitative variables were expressed as frequency and percentages.

## CURRENT STATE OF KNOWLEDGE

Our literature search identified 21 articles including 16 case reports and 5 case series covering a total of 31 cases of biliary hamartoma. All data concerning the demographics, clinical course and outcomes of these cases is shown in Table 1 [2-22].

**Table 1:**
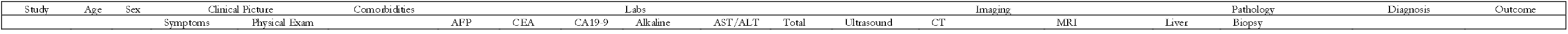
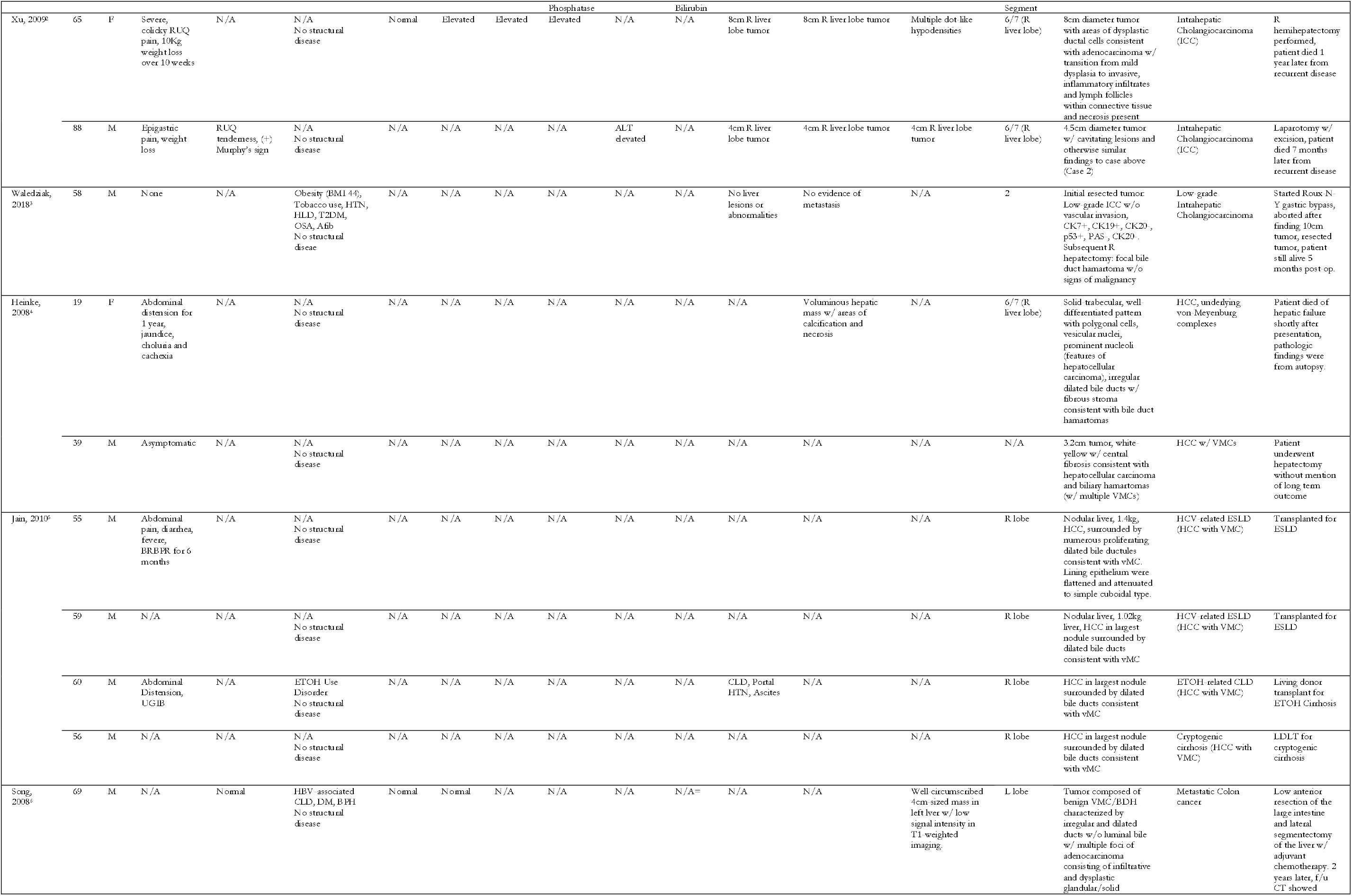
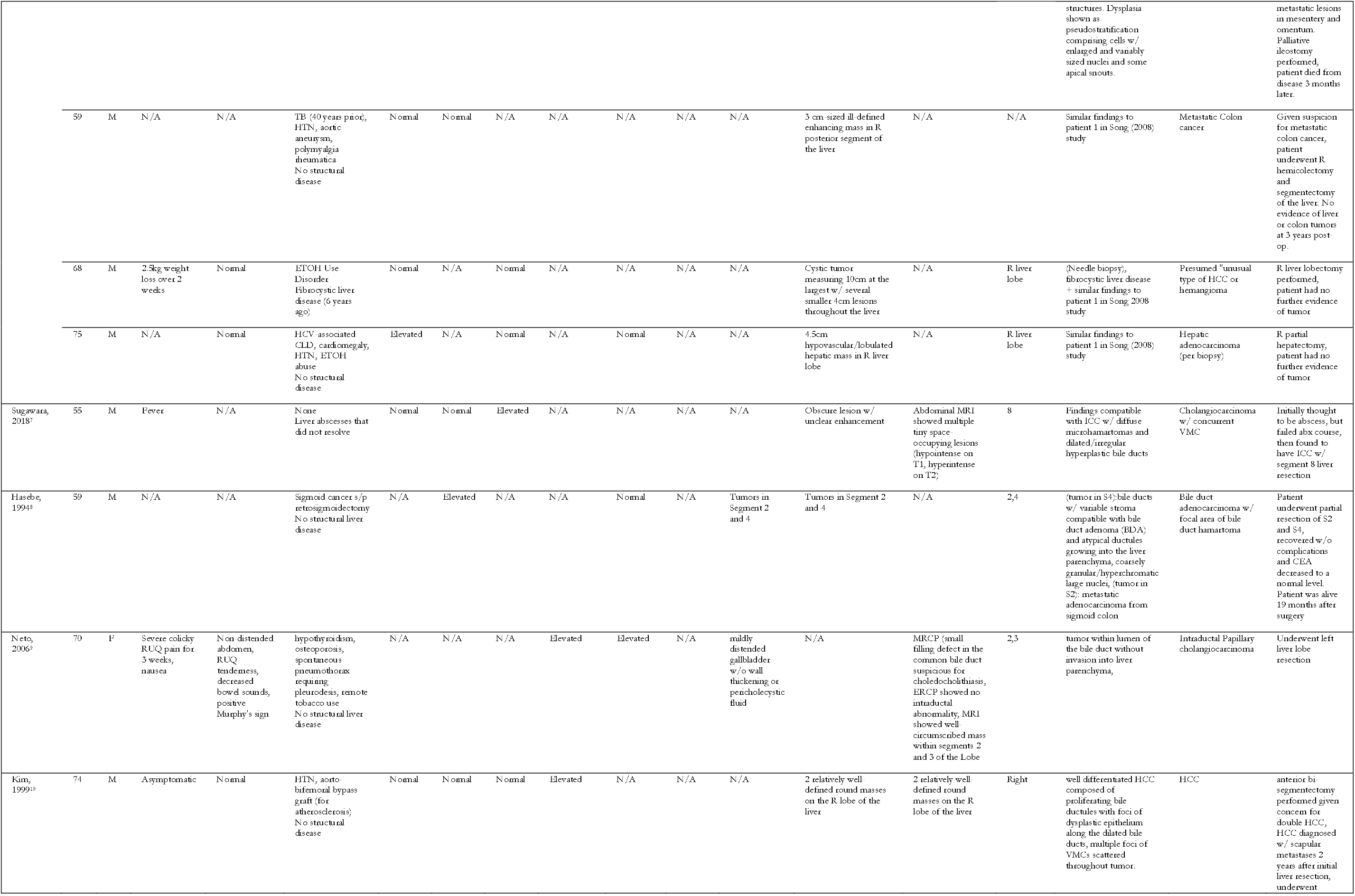
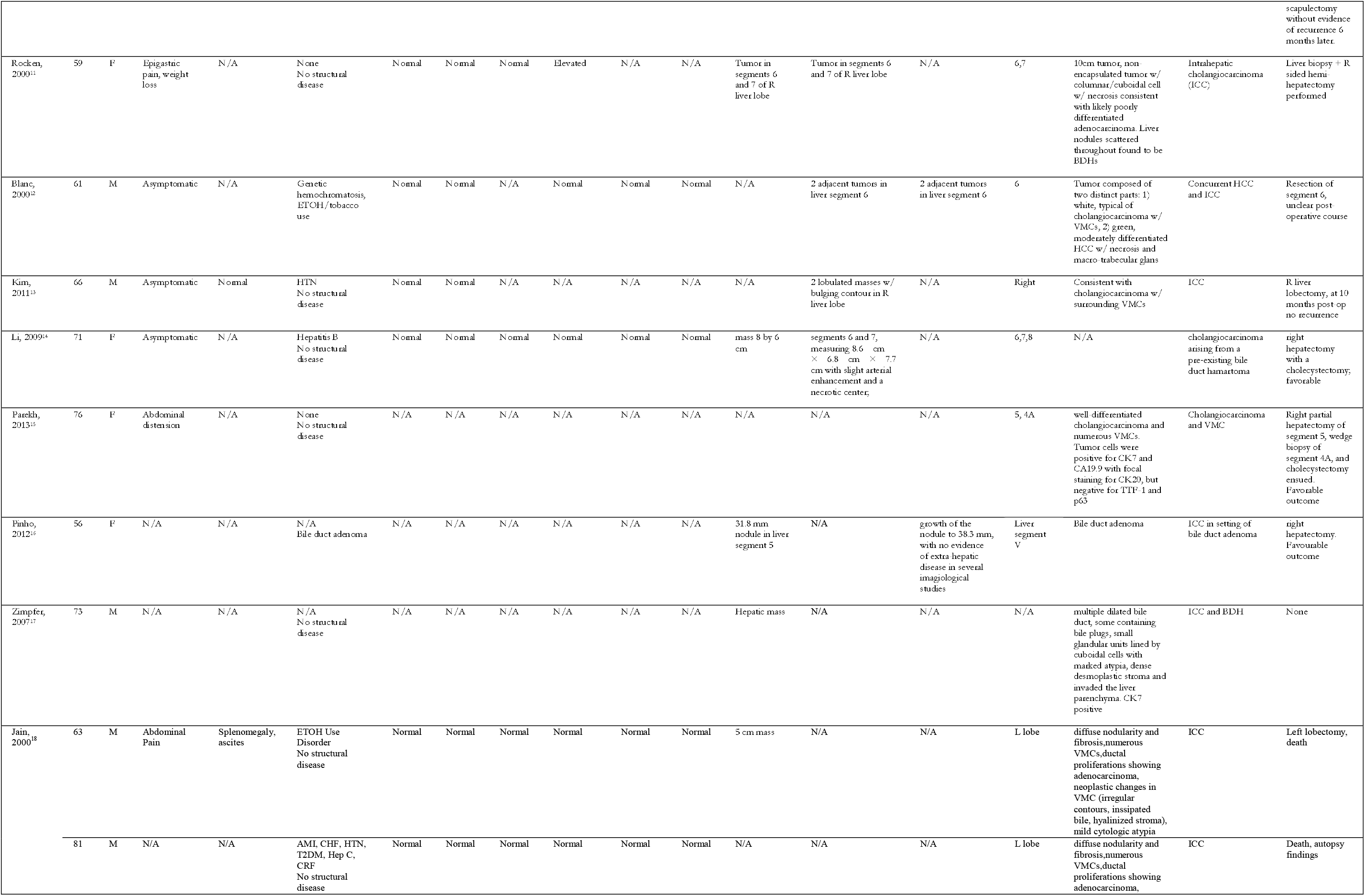
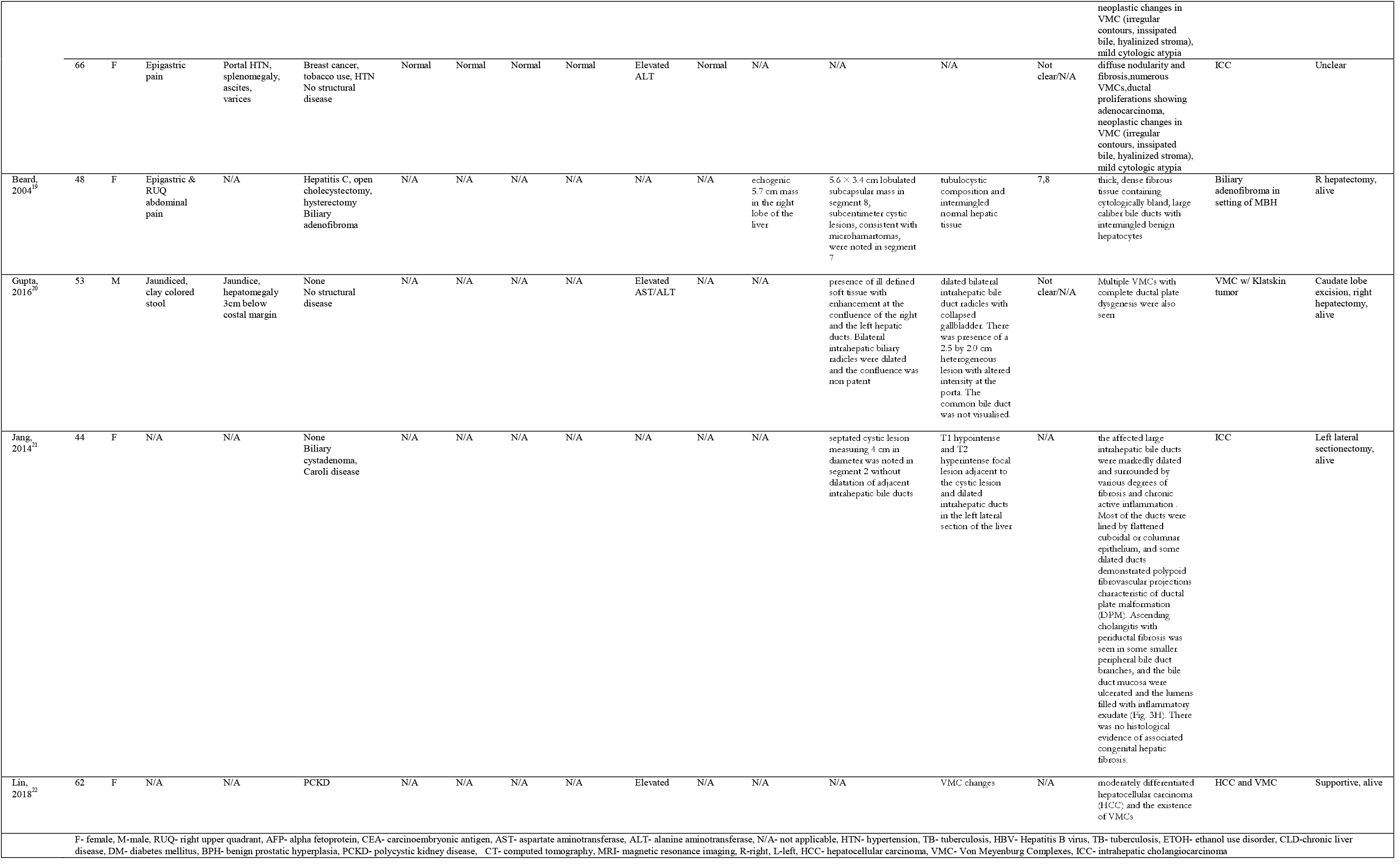
Summary of all the articles reviewed.

### Epidemiology

Cases included in this study covered a wide time period with the earliest case occurring in 1994. The mean age at onset was approximately 61 years±13 (min 19 to max 88). In regards to gender, cases were predominantly in male patients who made up 64.5% of the case population compared to 35.5% for females. Comorbidities were variably included in case descriptions with 54.8% of cases reporting comorbidities (17/31 cases). Of the comorbidities noted, there was no disproportionately high prevalence of one specific disease, although the most commonly reported were hypertension (22.6%), hepatitis C (16.1%) and alcohol use disorder (16.1%).

### Structural disease

There were six patients (19.4%) who had some form of previously diagnosed structural hepatobiliary disease. These diseases included fibrocystic liver disease (1/6 cases), polycystic liver and kidney disease (1/6 cases), biliary duct adenoma (1/6 cases), Caroli’s disease (1/6 cases), hemochromatosis (1/6 cases) and biliary adenofibroma (1/6 cases). The malignancies found in these specific cases included hepatocellular carcinoma (2/6 cases), hepatocellular carcinoma and cholangiocarcinoma (1/6 cases) and intrahepatic cholangiocarcinoma (1/6 cases).

### Clinical Picture

Of all reported cases, 41.9% reported symptoms (13/31 cases) with the most common symptom being abdominal pain (32.3%). All other reported symptoms which included weight loss, jaundice, fever, abdominal distension and upper GI bleed occurred in less than 10% of cases.

### Biochemical/Imaging Findings/Pathology

Of the 31 cases, biochemical data was reported in 21. Aspartate aminotransferase (AST) and alanine aminotransferase (ALT) were elevated in 19.4% of cases (6/21 cases). Alkaline phosphatase and GGT elevations occurred in 29.0% of cases (9/21 cases) and total bilirubin was elevated in 12.9% of cases (4/21 cases). Imaging modalities occurred variably with ultrasound being conducted in 12 cases, CT in 17 cases and MRI in 12 cases. Ultrasound findings were minimally reported in regards to echogenicity. Lesions were reported as hypointense on CT in 29% of cases (10/17). On MRI, lesions were reported as hypointense on T1-weighted MRI 29.0% of the time (vs hyperintense in 3.2%) and hypointense 9.7% of the time on T2-weight MRI (vs hyperintense 19.4% of the time). The size of the tumor was reported in 17 cases and was larger than 1 cm in 16/17 cases. The location of the lesion was reported in 25 cases and was most often found in the right liver lobe (35.5%) vs. the left liver lobe which occurred in 9.7% of cases. A specific liver segment was reported in the remaining 35.5% of cases. Biopsies were consistent with malignancy in 77.4% of cases (24/31 cases) and were reported as partially malignant with histology consistent with Von Meyenburg complexes in the remaining 22.6%. Most common reported associated malignancy was cholangiocarcinoma 54.8% (17/31), followed by hepatocellular carcinoma 29.0% (10/31), 2/31 cases were consistent with malignant colon cancer. 1 case had underlying hepatocellular and concomitant cholangiocarcinoma and 1 case had underlying biliary adenofibroma.

### Management/Outcomes

Management of the biliary hamartomas and associated malignancy was reported in 30 cases, 93.3% of which resulted in surgical intervention (28/30 cases). The patient died in the remaining 2 cases prior to surgical intervention. Outcomes were reported in 29 cases and 22.6% of patients died while the remaining 77.5% were alive or the outcome was unknown.

## DISCUSSION

Biliary duct hamartomas (BDH) also known as “von Meyenburg complexes’’ were first reported in 1918 [23]. These are small bile duct lesions, most often benign in nature, that are believed to be caused by abnormal embryologic development of the ductal plates [24].The lesions are often bile-filled and are found in both non-cirrhotic and cirrhotic livers [25]. Biliary duct hamartomas were originally reported in only 0.6 to 0.7% of surgical and autopsy liver specimens [26]. However a more accurate incidence of approximately 5.6% in adults and 0.9 % in children was reported by Redston in a study of 2843 autopsies [1]. On gross examination and imaging, BDH are frequently mistaken for metastatic tumors.

The risk of developing neoplasia in association with BDH appears to be small and the exact incidence is not known. Most of the reported cases with BDH malignant transformation are diagnosed in resected liver specimens or at autopsy. We suspect the incidence of malignancy and carcinoma associated with BDH may be higher and our systematic review is aimed at identifying the lab work, imaging findings, and common clinical characteristics including appearance, size, and location of the lesions that are possibly related to malignancy.

The direct relation between BDH and malignancy has been rarely observed in the literature. Our study covered 31 cases from 21 articles dating back to 1975 which showed BDH cases leading into a potential malignant transformation. BDH transforming into a malignant lesion is not a well-known hypothesis. The increased risk has been attributed to bile stasis and the subsequent interaction of the possible carcinogens present in bile with the parenchyma. The gradual transition from BDH to hyperplastic or adenomatous lesions and then cholangiocarcinoma has been reported [18] (Figure 2, 3). It has been postulated that bile exposure time is directly proportional to the risk of developing malignancy and therefore the chance of BDH converting into malignancy can be higher than other biliary lesions [18, 27, 28]. Supporting the premise, our review showed that the mean age at onset was 61 years. Our study showed a male predominance with 64.5% cases found in males. The gender patterns have shown to be varied by different sites in the hepatobiliary system. Gallbladder cancer is seen more in females and extrahepatic bile duct and ampulla of Vater cancers noted more in males corresponding to our findings [29].

**Figure 2:**
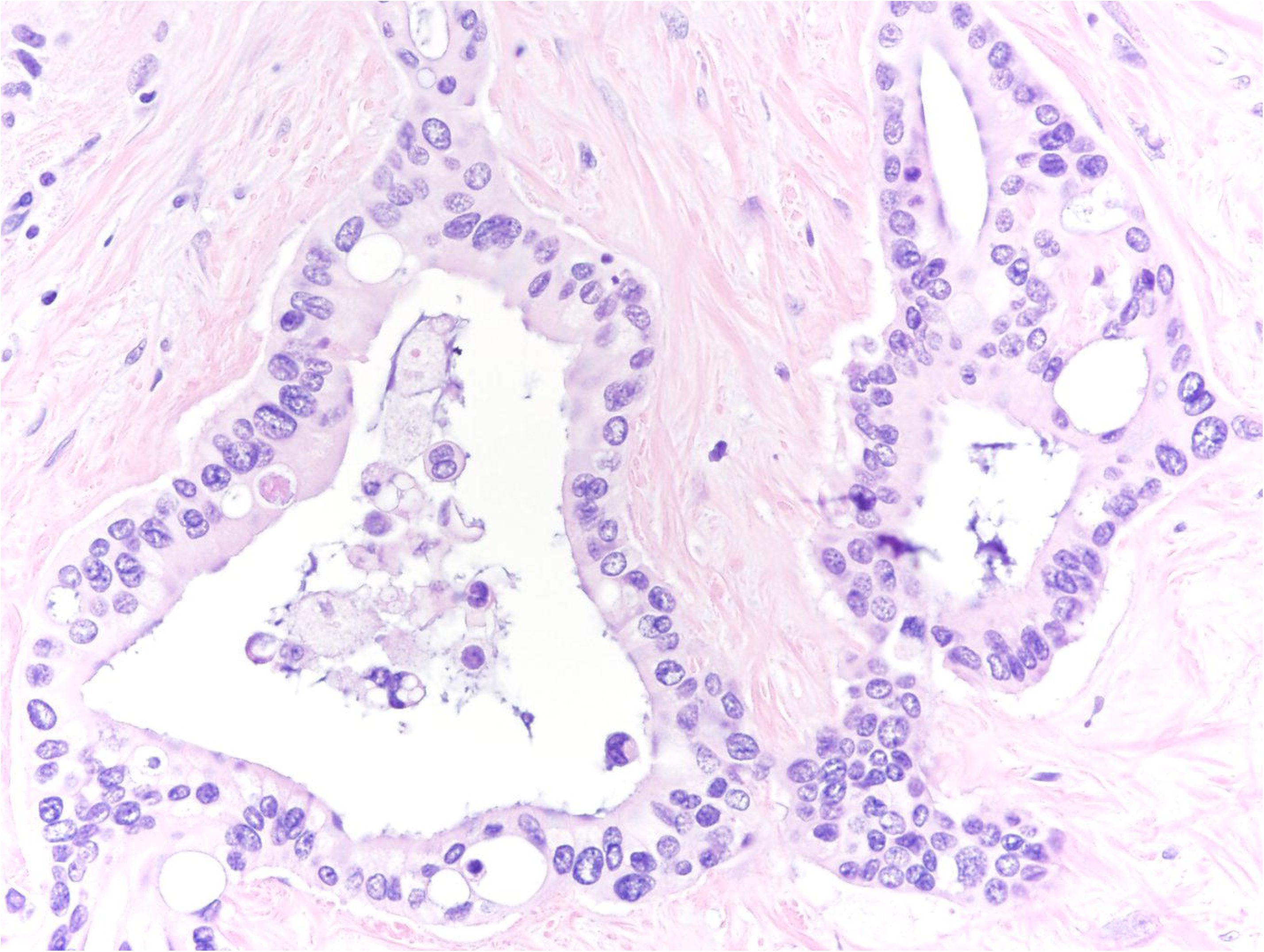
Liver parenchyma showing a well-demarcated lesion with numerous small to medium sized, irregularly shaped, angulated and dilated glands with intervening fibrous stroma consistent with biliary duct hamartomas. (H&E, x10)

**Figure 3:**
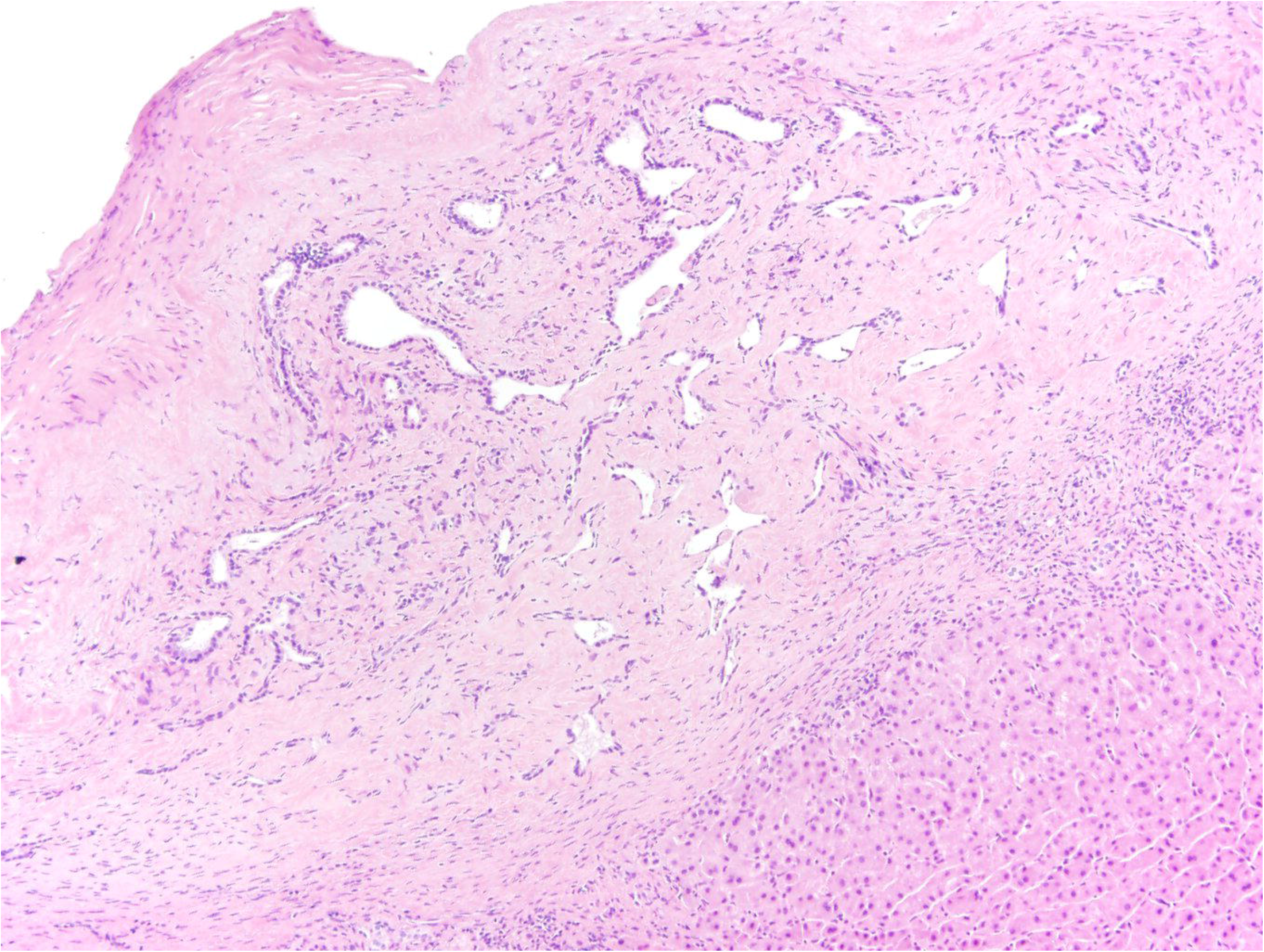
Well-differentiated irregular ductular to cribriform glands in a prominent fibrous stroma. The nuclei show mild to moderate atypia, are hyperchromatic and pleomorphic with nucleoli consistent with cholangiocarcinoma. (H&E, 40X)

As discussed earlier BDH and malignancy are rare and biliary hamartomas are mostly incidental findings at laparotomy and autopsy [24]. In our review, we made an attempt to find relations with BDH and different malignancies. The most common malignant tumors noted in the background of BDH are cholangiocarcinoma (CC), 17/31 cases, as reflected in our table followed by Hepatocellular carcinoma, 10/31. Histological review done by at least one of the studies revealed strong evidence of the evolution of BDH to CC through an intermediate stage of hyperplasia or adenomatous transformation and carcinoma-in-situ [18]. This is consistent with the current concept of a hyperplasia, dysplasia, neoplasia sequence seen in cholangiocarcinoma [30]. However, one study reported the prevalence of BDH as 5.6% in adults and suggested that combined with the lower number of cases, it should be assumed that both conditions may occur as a coincidence and not as a potential sequela [31].

HCC has also been observed in conjunction with BDH. Kim et al [13], reported a case with both HCC and CC in the presence of underlying BDH. In that study, no evidence was found of direct contact or histologic transition between HCC and CC, and only the CC had a transition area from the BDH as discussed above. Similarly in our review, all but 3 cases of HCC were associated with either viral illness, underlying cirrhosis, alcohol use, or concomitant CC. Perhaps it can be argued that these associations were the causative agents for HCC and BDH were just an incidental finding but the 3 cases reported by Heinke, and Lin make room for the debate as there were no precancerous factors associated with any of these cases [4, 22].

A few structural liver diseases also increase the risk of developing subsequent malignancies. This includes hemochromatosis. In a study conducted in 2011, 18.4% of the patients who had intrahepatic cholangiocarcinoma also had genetic hemochromatosis [32]. A similar finding was seen in a case report of a patient with polycystic liver and kidney disease [33]. Caroli’s disease is another structural problem that might increase the predisposition to developing malignancy [34]. Although, biliary duct hamartomas are a rare entity, it might be possible that another structural disease can increase the chances of developing dysplasia in a few benign lesions. In regards to clinical presentation, there were no clear clinical or biochemical findings that were present in a high proportion of cases that could be used as a predictor of disease onset. The most frequently reported symptom in this case series was abdominal pain (32.3% of cases), which is relatively non-specific. Laboratory abnormalities were also relatively non-specific as lab patterns consistent with cholestasis were only reported in 29.4% of cases.

Imaging findings were consistent with what is currently known about the radiographic characteristics of biliary hamartomas which is that they are typically 5-15mm, homogeneous lesions that appear hypointense on CT and T1-weighted MRI and hyperintense on T2-weighted MRI. Given the indolent, subclinical presentation of BDH, patients were generally not intervened upon until later in the disease course where surgical resection was often required.

There is variation in the sizes of these lesions on biopsy. According to one study, biliary duct hamartomas are about 0.2 cm in size, ranging from 0.1 to 0.4 cm whereas well-differentiated cholangiocarcinoma varies from 0.9 to 9.0 cm (mean: 4.3 cm) and moderate to poorly differentiated cholangiocarcinoma varies from 0.9 to 9.8 cm in terms of size [35]. The difference in sizes of biliary duct hamartomas and well-differentiated cholangiocarcinoma were significant implying that the size of the lesions might help to determine suspicion of malignancy. In our study, the majority of the malignant lesions were greater than 1 cm in size except for one lesion that was malignant despite being smaller than 1 cm suggesting that this discrepancy in size is an important factor. Although literature reports intrahepatic location (10.0%) of cholangiocarcinoma to be less frequent than the other locations [36], our study found that the majority (80.7%) of the suspicious lesions were located either in liver lobes or liver segments.

This case series has several limitations. The first being that it is retrospective, which limits the data available for analysis, as many of the cases did not report data such as lab findings and imaging. Case series are also subject to selection bias, as the studies included in the series are selected by individual researchers. Another limitation is the wide period of time between the earliest and the most recent study. As the earliest case was reported in the 1990s, imaging study selection would certainly be different between cases because MRIs would not be as commonly used in the early 1990s as compared to more recent cases. Perhaps the most important limitation is that there is limited data in the studies for immunohistochemistry results that could suggest or explain if cholangiocarcinoma may indeed occur in a pre-existing BDH.

In patients with either stable lesions or suspicion of biliary hamartomas, increasing symptoms, like worsening abdominal pain, weight loss, increase in size of lesions and obstructive jaundice would need further evaluation with possible biopsy to rule out underlying malignancy. Biliary hamartoma transformation to malignancy is under-reported but prevalence is still very low and in case of stable lesions, regular surveillance with imaging is not recommended and would not be a cost-effective approach.

## CONCLUSION

VMCs are an important entity when the question of malignancy potential comes into consideration. Patients with structural hepatobiliary diseases, chronic hepatitis, drug use and progressive symptoms might have an underlying hepatobiliary malignancy developing. This possibility of malignancy must be considered as a potential differential diagnosis when evaluating patients with worsening symptoms such as weight loss or increase in size of lesion. Regular surveillance is not recommended in patients with stable biliary hamartomas.

## What is known about this topic

- Biliary duct hamartomas are benign lesions usually found in surgical and autopsy liver specimens.

## What this study adds

- Patients with structural hepatobiliary disease or red flag symptoms such as weight loss, increase in size of lesion and obstructive jaundice should be evaluated for underlying malignant transformations.
- Biliary duct hamartomas are an important entity with malignancy potential which is under-reported.
- Regular surveillance is not recommended and not cost-effective in patients with stable lesions.

## Data Availability

Data may be made available on reasonable request from the corresponding author.

## Author Contributions

A.S. conceived the idea and contributed to the manuscript draft; A.N. contributed to the literature search; N.J. contributed to the data analysis; A.S., A.N., A.E.S., N.J., and K.L. wrote parts of the manuscript; A.S., provided the pathology slides; R.S. contributed to the supervision (attending), resources and critical review of the manuscript. All the authors have read and agreed to the final manuscript.

## Acknowledgements

The authors have no acknowledgments to declare.

## REFERENCES

1. Redston MS, Wanless IR. The hepatic von Meyenburg complex: prevalence and association with hepatic and renal cysts among 2843 autopsies. Mod Pathol. 1996; 9(3):233–237.

2. Xu AM, Xian ZH, Zhang SH, Chen XF. Intrahepatic cholangiocarcinoma arising in multiple bile duct hamartomas: report of two cases and review of the literature. Eur J Gastroenterol Hepatol. 2009; 21(5):580–584.

3. Walędziak M, Różańska-Walędziak A, Kowalewski PK, Paśnik K. Intrahepatic cholangiocarcinoma in an obese patient qualified for laparoscopic bariatric surgery - a case study. Wideochir Inne Tech Maloinwazyjne. 2018; 13(2):257–259.

4. Heinke T, Pellacani LB, Costa Hde O, Fuziy RA, Franco M. Hepatocellular carcinoma in association with bile duct hamartomas: report on 2 cases and review of the literature. Ann Diag Pathol. 2008; 12(3):208–211.

5. Jain D, Nayak NC, Saigal S. Hepatocellular carcinoma arising in association with von-Meyenburg’s complexes: an incidental finding or precursor lesions? A clinicopatholigic study of 4 cases. Ann Diagn Pathol. 2010; 14(5):317–320.

6. Song JS, Lee YJ, Kim KW, Huh J, Jang SJ, Yu E. Cholangiocarcinoma arising in von Meyenburg complexes: report of four cases. Pathol Int. 2008; 58(8):503–512.

7. Sugawara T, Shindoh J, Hoshi D, Hashimoto M. Intrahepatic cholangiocarcinoma and portal hypertension developing in a patient with multicystic biliary microhamartomas. Malays J Pathol. 2018; 40(3):331–335.

8. Hasebe T, Sakamoto M, Mukai K, Kawano N, Konishi M, Ryu M, et al. Cholangiocarcinoma arising in bile duct adenoma with focal area of bile duct hamartoma. Virchows Arch. 1995; 426(2):209–213. doi: 10.1007/BF00192644.

9. Neto AG, Dainiak C, Qin L, Salem RR, Jain D. Intraductal papillary cholangiocarcinoma associated with von Meyenberg complexes: a case report. Dig Dis Sci. 2007; 52(10):2643–2645.

10. Kim YW, Park YK, Park JH, Lee J, Lee SM, Hong SW, et al. A case with intrahepatic double cancer: hepatocellular carcinoma and cholangiocarcinoma associated with multiple von Meyenburg complexes. Yonsei Med J. 1999; 40(5): 506–509.

11. Röcken C, Pross M, Brucks U, Ridwelski K, Roessner A. Cholangiocarcinoma occurring in a liver with multiple bile duct hamartomas (von Meyenburg complexes). Arch Pathol Lab Med. 2000; 124(11):1704–1706.

12. Blanc JF, Bernard PH, Carles J, Le Bail B, Balabaud C, Bioulac-Sage P. Cholangiocarcinoma arising in Von Meyenburg complex associated with hepatocellular carcinoma in genetic haemochromatosis. Eur J Gastroenterol Hepatol. 2000; 12(2):233–237.

13. Kim HK, Jin SY. Cholangiocarcinoma arising in von Meyenburg complexes. Korean J Hepatol. 2011; 17(2):161–164.

14. Li WH, Tsang YW, Cheung MT. Cholangiocarcinoma arising from bile duct hamartoma. Surg Pract. 2009; 13(3): 83–85.

15. Parekh V, Reddy S, Peker D. Histologic Glimpse to Cholangiocarcinoma Arising in Von-Meyenburg Complexes: Report of a Rare Case. Am J Clin Pathol. 2013; 140 (Suppl 1): A107.

16. Pinho AC, Melo RB, Oliveira M, Almeida M, Lopes J, Graça L, et al. Adenoma-carcinoma sequence in intrahepatic cholangiocarcinoma. Int J Surg Case Rep. 2012; 3(4):131–133.

17. Zimpfer A, Nebel B, Terracciano L, Koch H. Intrahepatic cholangiocellular carcinoma associated with von Meyenburg complexes: case report and review of literature. Diagn Pathol. 2007; 2: S10.

18. Jain D, Sarode VR, Abdul-Karim FW, Homer R, Robert ME. Evidence for the neoplastic transformation of Von-Meyenburg complexes. Am J Surg Pathol. 2000; 24(8):1131–1139.

19. Beard RE, Yee EU, Mortele KJ, Khwaja K. Multicystic biliary hamartoma: A report of a rare entity and a review of the literature. Int J Surg Case Rep. 2014; 5(12):919–923.

20. Gupta A, Pattnaik B, Das A, Kaman L. Von Meyenburg complex and complete ductal plate malformation along with Klatskin tumour: a rare association. BMJ Case Rep. 2016; 2016:10.1136/bcr-2016-215220.

21. Jang MH, Lee YJ, Kim H. Intrahepatic cholangiocarcinoma arising in Caroli’s disease. Clin Mol Hepatol. 2014; 20(4):402–405.

22. Lin S, Shang TY, Wang MF, Lin J, Ye XJ, Zeng DW, et al. Polycystic kidney and hepatic disease 1 gene mutations in von Meyenburg complexes: Case report. World J Clin Cases. 2018; 6(9):296–300.

23. von Meyenburg H. Uber die Cystenleber. Beitr Pathol Anat. 1918; 64:477–532. In German.

24. Desmet VJ. Congenital diseases of intrahepatic bile ducts: variations on the theme ‘ductal plate malformation’. Hepatology. 1992; 16(4):1069–1083.

25. Torbenson MS. Hamartomas and malformations of the liver. Semin Diagn Pathol. 2019; 36(1):39–47.

26. Thommesen N. Biliary hamartomas (Von Meyenburg complexes) in liver needle biopsies. Acta Pathol Microbiol Scand A. 1978; 86(2):93–99.

27. Dekker A, Ten Kate FJW, Terpstra OT. Cholangiocarcinoma associated with multiple bile duct hamartomas of the liver. Dig Dis Sci. 1989; 34(6):952–958.

28. Scott J, Shousha S, Thomas HC, Sherlock S. Bile duct carcinoma: a late complication of congenital hepatic fibrosis. Am J Gastroenterol. 1980; 73(2):113–119.

29. Castro FA, Koshiol J, Hsing AW, Devesa SS. Biliary tract cancer incidence in the United States-Demographic and temporal variations by anatomic site. Int J Cancer. 2013; 133(7):1664–1671.

30. Anthony PP. Pathology of the Liver-3rd edition. 1994. New York. Churchill Livingstone.

31. Kendall T, Verheij J, Gaudio E, Evert M, Guido M, Goeppert B, et al. Anatomical, histomorphological and molecular classification of cholangiocarcinoma. Liver Int. 2019; 39 Suppl 1:7–18.

32. Sulpice L, Rayar M, Boucher E, Pele F, Pracht M, Meunier B. Intrahepatic cholangiocarcinoma: impact of genetic hemochromatosis on outcome and overall survival after surgical resection. J Surg Res. 2013; 180(1):56–61.

33. Sasaki M, Katayanagi K, Watanabe K, Takasawa K, Nakanuma Y. Intrahepatic cholangiocarcinoma arising in autosomal dominant polycystic kidney disease. Virchows Arch. 2002; 441(1):98–100.

34. Kasper HU, Stippel DL,Töx U, Drebber U, Dienes HP. Primäres Cholangiokarzinom auf dem Boden einer Caroli-Erkrankung. Fallbericht und Literaturübersicht [Primary cholangiocarcinoma in a case of Caroli’s disease: case report and literature review]. Pathologe. 2006; 27(4):300-304. In German.

35. Tsokos CG, Krings G, Yilmaz F, Ferrell LD, Gill RM. Proliferative index facilitates distinction between benign biliary lesions and intrahepatic cholangiocarcinoma. Hum Pathol. 2016; 57:61–67.

36. Razumilava N, Gores GJ. Cholangiocarcinoma. Lancet. 2014; 383 (9935):2168–2179.

